# Changes in the stool and oropharyngeal microbiome in obsessive-compulsive disorder

**DOI:** 10.1101/2020.05.26.20113779

**Authors:** Laura Domènech, Jesse Willis, Maria Alemany, Marta Morell, Eva Real, Geòrgia Escaramís, Sara Bertolín, Daniel Sánchez Chinchilla, Susanna Balcells, Cinto Segalàs, Xavier Estivill, Jose M Menchón, Toni Gabaldón, Pino Alonso, Raquel Rabionet

## Abstract

Although the etiology of obsessive-compulsive disorder (OCD) is largely unknown, it is accepted that OCD is a complex disorder. There is a known bi-directional interaction between the gut microbiome and brain activity. Several authors have reported associations between changes in gut microbiota and neuropsychiatric disorders, including depression or autism. Furthermore, a pediatric-onset neuropsychiatric OCD-related syndrome occurs after streptococcal infection, which might indicate that exposure to certain microbes could be involved in OCD susceptibility. However, only one study has investigated the microbiome of OCD patients to date. We performed 16S ribosomal RNA gene-based metagenomic sequencing to analyze the stool and oropharyngeal microbiome composition of 32 OCD cases and 32 age and gender matched controls. We estimated different α- and β-diversity measures and performed LEfSe and Wilcoxon tests to assess differences in bacterial distribution. OCD stool samples showed a trend towards lower bacterial α-diversity, as well as an increase of the relative abundance of *Rikenellaceae*, particularly of the genus *Alistipes*, and lower relative abundance of *Prevotellaceae*, and two genera within the *Lachnospiraceae: Agathobacer* and *Coprococcus*. However, we did not observe a different Bacteroidetes to Firmicutes ratio between OCD cases and controls. Analysis of the oropharyngeal microbiome composition showed a lower Fusobacteria to Actinobacteria ratio in OCD cases. In conclusion, we observed an imbalance in the gut and oropharyngeal microbiomes of OCD cases, including, in stool, an increase of bacteria from the *Rikenellaceae* family, associated with gut inflammation, and a decrease of bacteria from the *Coprococcus* genus, associated with DOPAC synthesis.

## INTRODUCTION

Obsessive-compulsive disorder (OCD) is a neuropsychiatric disorder characterized by intrusive and unwanted thoughts (termed obsessions) and repetitive behaviors or mental acts (called compulsions) that are performed to partially relieve the anxiety or distress caused by the obsessions. The etiology of OCD is largely unknown, although it likely involves a combination of genetic, neurobiological and environmental factors or events. Genetic association studies, including genome-wide association analysis and a multispecies approach integrating evolutionary and regulatory information, have highlighted genes involved in dopamine, serotonin and glutamate signaling, synaptic connectivity and the cortico-striato-thalamo-cortical circuit (CSTC) (Gasso et al., 2015; Mattheisen et al., 2015; Noh et al., 2017; Servaes et al., 2017; Stewart et al., 2013).

Several researchers have speculated that the microbiome might also play a role in the development of OCD (Rees, 2014; Turna et al., 2016). In fact, a recent pilot study of the gut microbiome in OCD has observed some dysbiosis in OCD cases compared to controls (Turna et al., 2020), and a previous work observed a difference in gut microbial composition in deer mice presenting an obsessive behavior compared to the normal individuals (Scheepers et al., 2019), while microbial treatments such as germ-free environment and probiotic treatments can modify OCD-like behavior in rodents (Bastiaanssen et al., 2019), and some OCD risk factors, such as stress, pregnancy or antibiotic use, are known to disrupt the gut microbiome (Rees, 2014). Moreover, it is recognized that some children, after suffering a streptococcal infection, present with a sudden onset of tics, OCD and other behavio**r**al symptoms, a condition known as pediatric autoimmune neuropsychiatric disorders associated with streptococcal infections syndrome (PANS/PANDAS) (Snider and Swedo, 2004).

There is a well-known bi-directional interaction between the gut microbiota and brain activity (Cryan and Dinan, 2012; Mayer et al., 2014). Brain function can be affected by endocrine-, neurocrine- and inflammation-related signals from the gut microbiota, while psychological and physical stressors can affect the composition of the gut microbiome (Bailey et al., 2011; Partrick et al., 2018). Differences in the composition of the gut microbiome have been associated to depression (Valles-Colomer et al., 2019), autistic disorder (Liu et al., 2019a; Tomova et al., 2015; Vuong and Hsiao, 2017), or schizophrenia (Schwarz et al., 2018), among other psychiatric disorders. Moreover, two studies have found differences in gut bacterial distribution when OCD cases or PANS/PANDAS patients are compared to healthy age-matched individuals (Quagliariello et al., 2018; Turna et al., 2020). On the other hand, some studies have also shown an alteration of the oropharyngeal microbiome in patients with autism (Hicks et al., 2018; Qiao et al., 2018) or schizophrenia (Castro-Nallar et al., 2015). However, so far, only one study has explored a potential link between gut microbiota and OCD, and the OCD oral microbiota has not been explored yet. Here, we explore the composition of fecal and oropharyngeal microbiota of OCD-patients before and after treatment and compare it to healthy matched controls.

## MATERIAL AND METHODS

### Sample collection

Thirty-eight patients (20 females; mean age=40.16±14.12, see Table 1) with a diagnosis of OCD with at least one year of development were recruited from the OCD clinic at Bellvitge Hospital (Barcelona, Spain) from January 2016 to September 2017. Diagnoses were assigned by two psychiatrists with extensive clinical experience in OCD, following the DSM-IV criteria for OCD diagnosis (Association, 1994) and using the Structured Clinical Interview for DSM-IV Axis I Disorders-Clinician Version (SCID-I) (First et al., 1996). A summary of the patients’ clinical data is provided in supplementary Table S1. Patients presenting psychoactive substance abuse/dependence (current or in the past six months), psychotic disorders, intellectual disability, severe organic or neurological pathology (excepting tic disorder), or autism spectrum disorder were excluded from the study. Other affective and anxiety disorders were not criteria for exclusion in cases where OCD was the main diagnosis.

**Table 1.** Age and gender characteristics for the control and OCD sample cohorts.

| Group | Female | Male | Age | Diet |  |  |  |  |  |
| --- | --- | --- | --- | --- | --- | --- | --- | --- | --- |
|  |  |  | Aver (SD; range) | Q6 | Q7 | Q11 | Q12 | Q17 | Q18 |
| OCD | 20 | 18 | 40.16 (14.12;18-71) | 1.5 | 2.4 | 3.1 | 2.2 | 2.1 | 2.5 |
| Control | 18 | 15 | 36.00 (9.87; 23-59) | 2.15 | 2.4 | 3.6 | 2.1 | 2.1 | 2.5 |

Female/Male: number of female/male participants; Aver: average; SD: standard deviation; Diet: average value for group responses to questionnaire questions Q6 (alcohol); Q7 (yoghurt, kefir...); Q11 (vegetables); Q12 (beans); Q17 (red meat) and Q18 (white meat); values range from 1 (not in last 7 days) to 4 (daily). For more details, see questionnaire in supplementary table S3.

Thirty-three healthy controls (18 females; mean age=36±9.87) were recruited from the same sociodemographic environment and constituted a comparable sample in terms of age and gender (see Table 1 and supplementary Table S2 for a summary of clinical information on controls). Prior to inclusion, each control participant underwent the Structured Clinical Interview for DSM-IV non-patient version (SCID-NP) (First et al., 2002) to exclude presence or history of any psychiatric disorder.

Written informed consent was obtained from all participants after a complete description of the study, which was performed in accordance with the Declaration of Helsinki (2013) and approved by Bellvitge Hospital’s ethical committee.

Stool samples from 28 OCD cases at two time-points (before (OCD T0) and after (OCD T3) three months of pharmacological treatment and cognitive behavioral therapy), 7 OCD cases at a single time-point (4 OCD T0 and 3 OCD T3), and 33 healthy subjects were collected in Stool Collection Tubes (Stratec Molecular). Samples were stored at −20°C until processed. DNA was extracted using the PSP Spin Stool DNA Basic Kit (Stratec Molecular).

Oropharyngeal swab samples, corresponding to the tonsils and the back of the throat, from 28 OCD cases at two time-points, 8 single timepoint OCD cases (4 OCD T0 and 4 OCD T3) and 32 healthy individuals were collected with the Catch-All sample collection swabs (Epicentre), conserved in PowerBead tubes (MO BIO Laboratories), frozen at −20°C and processed for DNA extraction with the PowerSoil DNA Isolation kit (MO BIO Laboratories). Details for the overlap between stool T0, stool T3, oropharyngeal T0 and oropharyngeal T3 samples are provided in Figure S1.

Sample sizes were deemed appropriate for this exploratory approach considering previous similar studies of microbiota involvement in other psychiatric disorders (Castro-Nallar et al., 2015; Qiao et al., 2018; Quagliariello et al., 2018; Schwarz et al., 2018; Tomova et al., 2015; Turna et al., 2020), performed with similar sample sizes.

All participants were asked to provide dietary information through a questionnaire (supplementary tables S3 and S4). Severity of the obsessive and compulsive symptoms before and after treatment among the OCD patients was assessed through the clinician-administered version of the Yale-Brown Obsessive Compulsive Scale (Y-BOCS) (Goodman et al., 1989). A global measure as well as independent ones for both obsessions and compulsions were taken. Table 1 and supplementary Tables S1 and S2 summarize the main characteristics of the OCD and control cohorts.

### Sequencing and data analysis

16S ribosomal RNA (rRNA) sequencing was performed by the UPF Genomics Core Facility following Illumina’s protocol with slight modifications. Briefly, in each sample, the variable V3-V4 region of the 16S rRNA gene was amplified for 35 cycles using the primers described in Klindworth et al. (Klindworth et al., 2013) and Platinum High Fidelity Taq (Thermofisher) and purified, followed by 8 cycles of indexing PCR, and quantification. The indexed products were then pooled in an equimolar way in two final amplicon libraries (one for stool and one for tonsils), each containing 96 samples. Each library was sequenced in one 2 × 300 bp paired-end sequencing run on a MiSeq System.

The DADA2 R package (version 1.6.0) (Callahan et al., 2016) was employed to obtain counts of amplicon sequence variants (ASV), and sequence taxonomy was assigned using the SILVA database (Yilmaz et al., 2014). Resulting ASV counts, along with clinical data and diet information collected for each individual, were stored and analyzed using the Phyloseq package (version 1.22.3). 16S counts were normalized per sample, obtaining the relative abundance of each taxon within a sample, with all values ranging between 0 and 100.

We estimated different α- (within samples) and β-diversity (between samples) measures using the Phyloseq, picante and vegan R packages (see supplementary material for details). α-diversity indices included the Shannon and Simpson Diversity Index, Faith’s phylogenetic diversity, the observed species index, the Chao1 index and the ACE (Abundance-based Coverage Estimator) index. β-diversity measures included Unifrac distances, the Bray-Curtis dissimilarity, Jensen-Shannon diversity and Canberra indices. We performed a PERMANOVA test on β-diversity with 999 permutations considering even dependence of samples (paired OCD samples after and before treatment). Finally, we applied a Principal Coordinate Analysis (PCoA) to visualize the clustering of the samples.

### Statistical analysis

Correlation between categorical variables (i.e. sample type, type of obsessions, etc.) and taxa abundances or other continuous variables was tested using the Kruskal-Wallis rank sum test, and correlation between two categorical variables was tested using Chi-squared tests. Statistical significance of α-diversity differences between groups was evaluated with Mann–Whitney U test when samples were independent, and with Wilcoxon rank-sum test when samples were paired. In all cases, we applied the Bonferroni correction to adjust the p-values by the number of comparisons and provide only corrected p-values. Correlation plots and boxplots were generated using *ggplot2* (version 2.2.1) and association plots were generated using the *assoc* function from the R package *vcd* (version 1.4.4).

Linear discriminant analysis Effect Size (LEfSe) was performed using the Huttenhower lab’s tool implemented in Galaxy web (Segata et al., 2011). For the statistical test we used an α value of 0.05 and a logarithmic LDA score threshold of 2.0.

## RESULTS

High-throughput sequencing analysis of bacterial 16S rRNA V3-V4 regions was conducted on fecal and oropharyngeal samples of the 32 OCD (two timepoints) and 33 control individuals (see Figure S1). In total, 8,653,960 high-quality reads (90,145.42 ± 15,356.44 reads/sample) were obtained from all 96 fecal samples, representing 196 operational taxonomic units (OTUs) when considering all samples, while each sample had, on average, 91.77 ± 17.51 OTUs. For the 96 oropharyngeal samples, 6,051,021 high-quality reads (63,031.47 ± 16555 reads/sample) were obtained, representing 81 operational taxonomic units for all samples, with an average of 32.22 ± 9.29 OTUs/sample.

While statistical analyses revealed some differences between cases (at T0 and/or at T3) and controls, no statistically significant differences were identified when comparing OCD cases before and after treatment (T0 vs T3). On the other hand, we observed some association between some of the diet variables and taxa abundances, but there were no significant associations between the variables collected in the diet questionnaires and whether the individual belonged to the OCD or control group, which suggests that the taxa differences between OCD cases and controls may be influenced by the OCD phenotype, rather than by other variables.

### OCD samples show a trend towards lower bacterial α- diversity in the gut

Analysis of the microbial composition of the gut showed that OCD T0 samples presented an overall lower level of all α-diversity indices compared to the control group, although no test reached statistical significance after adjusting for multiple testing (Bonferroni adjusted p-value = 0.057) (Figure 1A). These differences were reduced in the OCD T3 group. On the other hand, we did not observe a strong separation of sample groups based on the β-diversity measures (Figure S2), and there were no statistical differences between the Bacteroidetes to Firmicutes ratio when comparing OCD samples to controls (Wilcoxon rank-sum test Bonferroni adjusted p-value = 0.0653; Figure 2A). In all three groups, the 20 most abundant genera belonged to either the Bacteroidetes or Firmicutes Phyla, while at the family level there was also representation of *Proteobacteria, Verrucomicrobia* and *Actinobacteria*, albeit at very low abundances (Figure S3).

**Figure 1.**
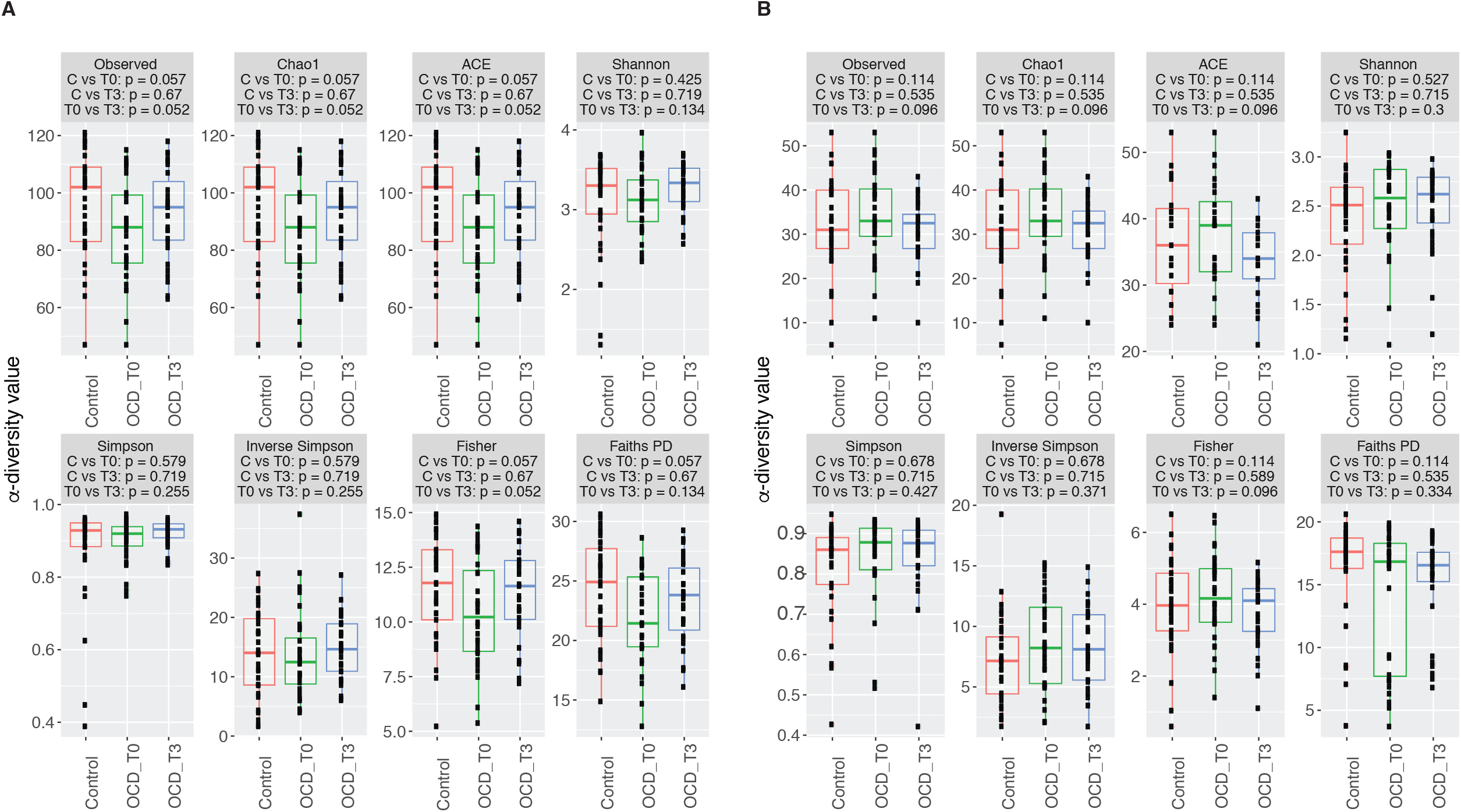
Boxplots representing α-diversity indices: Observed, Chao1, ACE, Shannon, Simpson, Inverted Simpson, Fisher and PD. The plots represent the median, 25th, and 75th percentiles calculated for Controls (red), OCD T0 (green) and OCD T3 (blue) in stool samples (**a**) or oropharyngeal samples (**b**). The corresponding Bonferroni adjusted p-values are reported below each index (OCD T0 vs controls and OCD T3 vs controls was calculated with Mann–Whitney U test; OCD T0 vs 0CD T3 with Wilcoxon rank-sum test).

**Figure 2.**
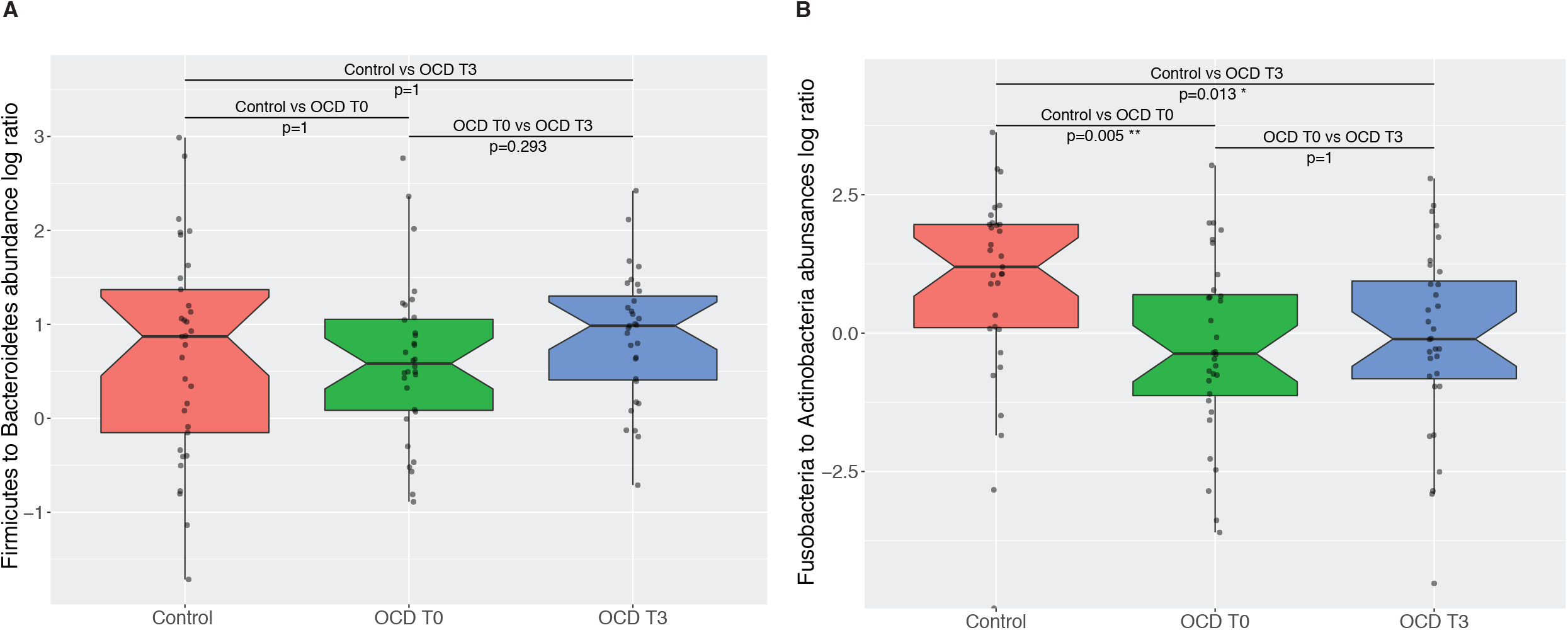
Notched boxplots representing the values of the ratio for the two most abundant taxa in each area: **a**. Firmicutes to Bacteroidetes ratio in gut samples from Control, OCD T0 and OCD T3 subjects. **b**. Fusobacteria to Actinobacteria ratio in Oropharyngeal samples from Control, OCD T0 and OCD T3 subjects. Bonferroni adjusted p-value of Wilcoxon rank-sum test between Controls and OCD T0, and controls and OCD T3 are indicated.

### LEfSe results reveal an enrichment for *Rikenellaceae* and depletion of *Prevotellaceae*, and a shift from *Coprococcus* to *Flavonifractor* within the Clostridiales in the gut

LEfSe analysis revealed a significant increase of the relative abundance of *Rikenellaceae*, particularly of the *Alistipes* genus, and a decrease in the levels of *Prevotellaceae*, in OCD T0 stool samples compared to controls (Figure 3). We also observed a different distribution of bacteria from the order Clostridiales, where OCD T0 samples presented a higher abundance of the genera *Oscillibacter, Anaerostipes* and *Flavonifractor*, and a depletion of *Agathobacter, Coprococcus, Lachnospira, Howardella, Romboutsia, Butyricicoccus* and *Clostridium* compared to controls. In addition, within these, depletion of *Lachnospira pectinoschiza* correlated with OCD severity on the obsessions subscale (YBOCS_OBS_0 vs Species *Lachnospira pectinoschiza r* =-0.420). Relative abundances in stool samples from cases and controls for these taxa are presented in Table S5. Most of these differences are also present when comparing OCD post-treatment samples versus controls (Figure S4).

**Figure 3.**
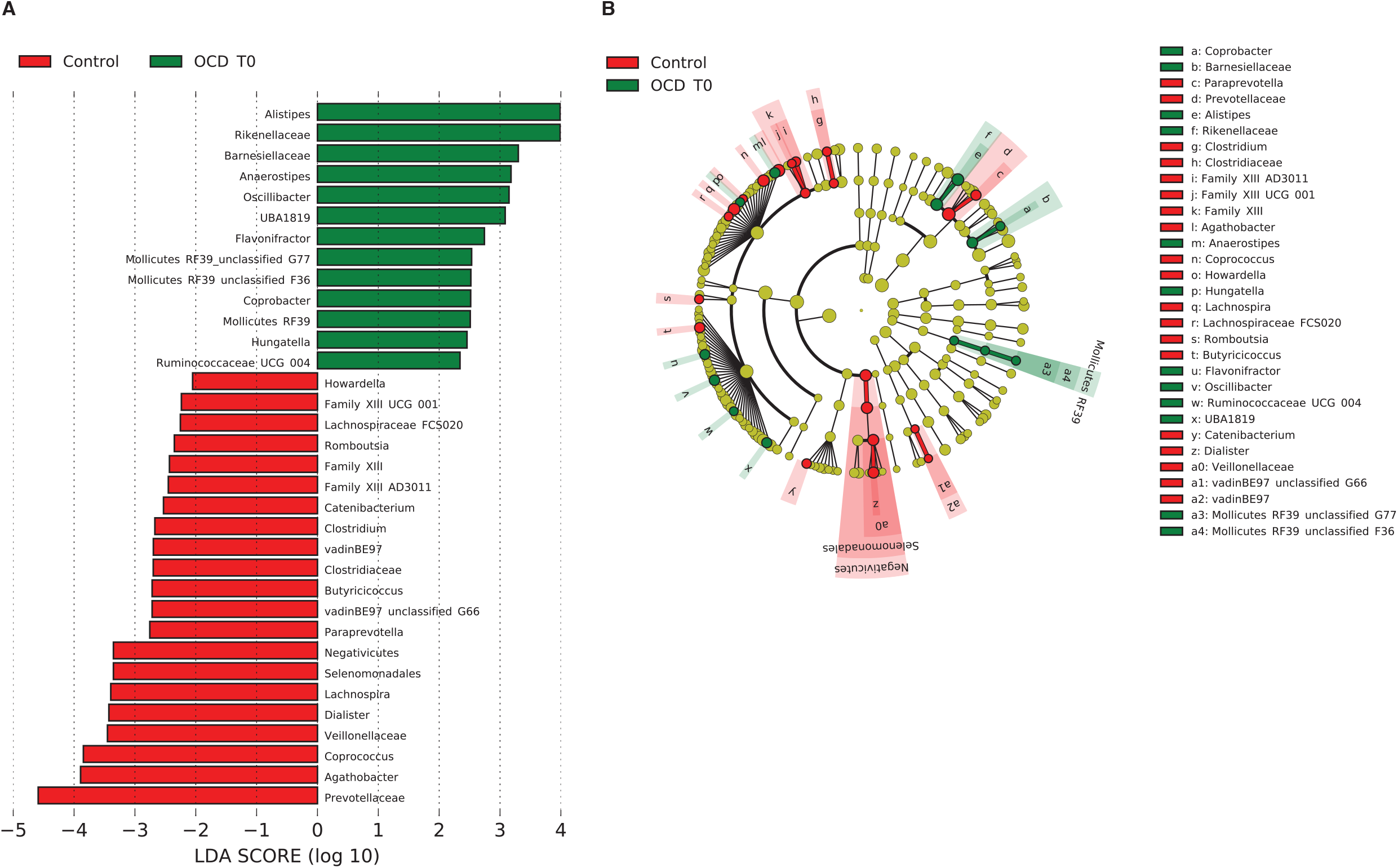
**a**. Biomarkers associated with OCD and control groups discovered by LEfSe analysis (α value=0.05, logarithmic LDA score threshold=2.0) in stool samples. **b**. Cladogram representing the phylogenetic relationship of biomarkers associated with OCD and control groups through the Linear discriminant Effect Size (LEfSe) analysis (α value=0.05, logarithmic LDA score threshold=2.0) in stool samples.

### In the oropharyngeal microbiome, OCD samples show a lower Fusobacteria to Actinobacteria ratio

Again, in the analysis of the oropharyngeal microbiome, α-diversity indices did not show significant differences between the OCD and control groups (Figure 1B). Similarly, we did not see any separation of sample groups based on most β-diversity measures (Figure S5). However, there was a significant difference in the ratio of Fusobacteria to Actinobacteria when comparing OCD samples (T0 and T3) to controls (Wilcoxon rank-sum test p-value = 0.004; c, Figure 2B), where OCD samples had lower ratios. Bacterial diversity in the oropharyngeal samples was lower than in the stool samples (Shannon diversity: Stool= 3.17 ± 0.46, oropharyngeal= 2.46 ± 0.45; Faiths PD: Stool= 23.18 ± 4.00, oropharyngeal= 15.25 ± 4.53). Among the 20 most abundant genera, the largest contribution belonged to the Actinobacteria and Fusobacteria phyla, although Epsilonbacteraeota (e.g. *Campylobacter)*, Bacteroidetes, Firmicutes and Proteobacteria were also represented (Figure S6).

LEfSe analysis of the oropharyngeal microbiome data revealed an increase of species within the Actinobacteria and Coriobacteriia classes in the OCD samples: particularly, the genera *Actinomyces* and *Atopobium* represented a higher percentage of total bacteria in OCD T0 compared to control samples; there was also an increase of *Lachnospiraceae* and other Firmicutes. In contrast, control samples had a higher percentage of bacteria from the Fusobacteriia class (Figure 4). Analysis of OCD T3 vs controls highlighted again the increase of the Actinobacteria and Coriobacteriia (Figure S7). Relative abundances in oropharyngeal samples from cases and controls for these taxa are presented in Table S6.

**Figure 4.**
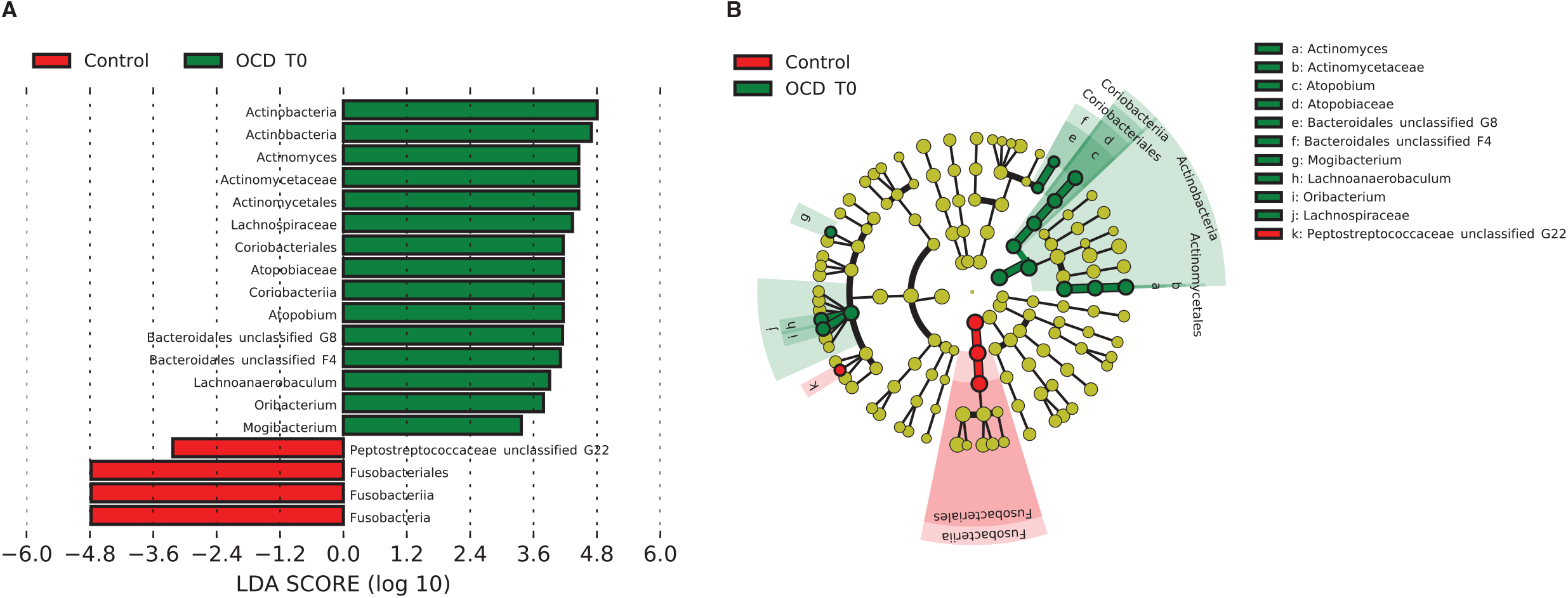
**a**. Biomarkers associated with OCD and control groups discovered by LEfSe analysis (α value=0.05, logarithmic LDA score threshold=2.0) in oropharyngeal samples. **b**. Cladogram representing the phylogenetic relationship of biomarkers associated with OCD and control groups through the Linear discriminant Effect Size (LEfSe) analysis (α value=0.05, logarithmic LDA score threshold=2.0) in oropharyngeal samples.

We also performed a directed analysis of enrichment of *Streptococcus* in OCD samples versus controls, as this genus has been shown to be associated to the onset of PANDAS (Quagliariello et al., 2018). However, the top 15 most abundant species did not include bacteria form this genus in either OCD T0, OCD T3, or controls.

## DISCUSSION

We have performed an explorative comparative analysis of the gut (fecal) and oropharyngeal microbiome between OCD cases (before and after treatment) and age- and gender-matched healthy controls, via 16S rRNA amplicon sequencing.

While our analysis shows only a trend towards lower bacterial diversity in the gut of OCD patients which is not significant after correction for multiple testing, this result is consistent with the lower gut α-diversity in PANS/PANDAS patients reported by Quagliariello et al. (2018), and in OCD by Turna et al. (2020). Lower α-diversity has also been reported in subjects with attention deficit hyperactivity disorder (Prehn-Kristensen et al., 2018), and in some studies of ASD individuals (Kang et al., 2018; Kang et al., 2013; Liu et al., 2019b), although it is not consistently observed, and may be dependent on the cohort characteristics, including sample size. Nevertheless, most of these studies, as ours, are exploratory analyses with small sample sizes, which may explain differences in the specific indexes reported significant. In addition, in the analysis of Quagliariello et al. (2018) they also observed, in the younger subset of PANS/PANDAS patients (aged 4-8 years), a higher percentage of Bacteroidetes and lower level of Firmicutes, while we did not observe significant shifts in the Bacteroidetes to Firmicutes ratio. This discrepancy between our results and those of Quagliariello et al. (2018) could be related to the age of our subjects, as all our patients are adults, while their study was performed on pediatric cases. However, we did not observe a correlation between age and any of the taxa ratios tested. Additional analyses on larger cohorts should clarify the observed trends.

The Wilcoxon and LEfSe tests performed here highlighted differences in OTU abundances in the gut: interestingly, we observed an increase of bacteria from the *Rikenellaceae* family, which had been reported as a biomarker of PANDAS (Quagliariello et al., 2018). In addition, an increase in abundance of *Rikenellaceae* has also been reported in ADHD (Aarts et al., 2017) and major depressive disorder (MDD) (Jiang et al., 2015). This bacterial family and, specifically, the *Alistipes* genus, is also positively associated with gut inflammation in human and mice studies (Bassett et al., 2015; Saulnier et al., 2011). Recently, a study linked neuroinflammation throughout the CSTC circuit to OCD (Attwells et al., 2017). Considering these observations, it is tempting to speculate a possible link between the neuroinflammation observed in OCD patients and the higher abundance *of Alistipes and Rikenellaceae* in these patients. Finally, OCD T0 samples showed lower levels of bacteria of the *Prevotellaceae* family, similarly to reports on ASD (Liu et al., 2019b).

LEfSe analysis also found specific members of the Firmicutes phylum with higher abundance in OCD T0 samples: *Oscillibacter, Anaerostipes*, and *Flavonifractor*, and a lower abundance *of Coprococcus and Lachnospira;* futhermore, abundance of *Lachnospira pectinoschiza* was also negatively correlated with severity, in line with the depletion in cases compared to controls. While several of these differences are maintained when comparing OCD post-treatment samples to the control group, the decrease in the genus *Lachnospira* is not. Furthermore, Valles-Colomer et al. (2019) report a correlation of four mental well-being scores (measured by the RAND-36 health-related quality of life survey) with higher abundance of *Coprococcus*, and two of these (social functioning and emotional well-being) also correlate with a lower abundance of *Flavonifractor* genus in the gut microbiota. We observe precisely the opposite trend in our cohort of OCD patients, where OCD patients have lower abundance of *Coprococcus* and higher abundance of *Flavonifractor*. The relative abundance of *Coprococcus* in the aforementioned study was strongly associated with the microbial pathway for DOPAC (a dopamine metabolite) synthesis. Relevantly, dopaminergic transmission has been implicated in the neurobiology of OCD, as dopamine receptor agonists are effective as an augmentation therapy to selective serotonin reuptake inhibitors in reducing OC symptoms (Vulink et al., 2009), and OCD patients under deep brain stimulation show improvements associated with release of striatal dopamine (Figee et al., 2014). However, our results are not replicating the findings by Turna et al. (2020). In their study, they observed a lower relative abundance of *Oscillospira, Odoribacter* and *Anaerostipes*. We do not see a significant difference for *Oscillospira* and *Odoribacter*, although a detailed look at the data show that, for *Oscillospira*, a similar trend is present, with lower abundance in OCD T0 samples compared to controls, and an increase in abundance in OCD T3 samples (supplementary table s5, figure S8). On the other hand, in the case of *Anaerostipes* we observe an increase of these bacteria as opposed to what was previously reported. These differences might be explained by chance, as both studies have a small, exploratory, sample size, or it could be related to dietary differences or to methodological differences in the analysis between both studies.

We also analyzed the oropharyngeal microbiome of OCD cases and controls. While the composition of the oral microbiome has received little attention in relation with psychiatric diseases, there are some studies that indicate possible changes in the oral microbiota of Parkinson’s disease or autistic disorder (Clarke et al., 1998; Hicks et al., 2018; Mihaila et al., 2019). In addition, the bacterial composition of the oral cavity might be used to represent the composition of the upper gastrointestinal tract (Tsuda et al., 2015). Furthermore, in this case we were particularly interested in the evaluation of the presence of *Streptococcus pyogenes*, as a potential biomarker of OCD, as it had been associated with PANDAS (Orefici et al., 2016), but we did not detect this species in our dataset. We then analyzed bacterial diversity measures between cases and controls without observing an overall difference in α-diversity. We did observe a significantly higher Actinobacteria to Fusobacteria ratio in OCD T0 compared to controls, which is still present, but reduced, after treatment. This difference correlated with an increase in *Actinomycetales* and a decrease of *Fusobacteriales* observed in the LEfSe analysis. We then analyzed whether the Fusobacteria to Actinobacteria ratio could be used as a proxy for the increased abundance of *Alistipes* and *Rikenellaceae* in the gut microbiome, but we did not detect a significant correlation with either *Alistipes* or *Rikenellaceae*. The Fusobacteria/Actinobacteria ratio was, however, correlated with the presence of the *Coprococcus* genus or the *Eggerthellaceae* family (Figure S9).

While our results support an imbalance in OCD cases, both in the gut and the oropharyngeal microbiome, it is not possible to establish a causal relationship, as the observed changes in the microbiome can be both a cause or a consequence of the disorder. It has been shown that the brain can modulate the gut microbiome by a top-down function of the gut-brain axis. For instance, the lower α- diversity observed in OCD cases could be a consequence of the anxiety provoked by obsessions. Since our study included only patients already diagnosed with OCD, it is not possible to discern between cause and effect. Further studies involving animal models, larger sample sizes, longitudinal cohorts and eventually, interventional experiments, should help understand the contribution of intestinal and oral microbiota to obsessive-compulsive disorder. In this regard, a recent animal study points toward the existence of an underlying aetiological association between alterations in gut microbiota and development of obsessive-compulsive behavior (Scheepers et al., 2019). Our analysis has some limitations: the study sample size is relatively small, although in line with other exploratory studies in autism or depression; on the other hand, gut microbiome composition has been analyzed through the bacterial composition of stool samples, which represents mostly the composition of the colon, while underestimating that of other regions of the gut such as the small intestine. Nevertheless, most studies on the gut microbiome use this same sample source, as it is more accessible and less invasive.

In summary, our results and those of others indicate that the gut microbiome might be playing a role in the pathogenesis of OCD, which might involve an increase in inflammation levels related to overabundance of bacteria in the *Alistipes* genus, or be related to a decrease in abundance of *Prevotella* and *Coprococcus* genera, the latter associated with the microbial DOPAC synthesis pathway. While awaiting confirmation on additional cohorts, these results open the door to further explorations into possible interventions directed to the modulation of both the microbiota in OCD patients or the mechanisms through which they exert their effect.

## Data Availability

NGS sequencing data is available upon request, and will be made available through the EGA data server.

## ACKNOWLEDGEMENTS

This project was supported by grants from the Spanish ministry of science and innovation (MICINN; SAF2013-49108-R); the Carlos III Health Institute (PI16/00950 PI18/00856); FEDER funds (‘A way to build Europe’) and by the Agency of University and Research Funding Management of the Catalan Government (2014SGR1672). LD was supported by a Severo Ochoa grant (SVP-2013-068066), MA was supported by the Secretariat for Universities and Research of the Ministry of Business and Knowledge of the Government of Catalonia Grant co-funded by the European Social Fund (ESF) “ESF, Investing in your future” (2017 FI_B 00327), DSC was supported by a grant from the MICINN (BES-2014-069814) and RR was supported by a fellowship from the Health Department of the Generalitat de Catalunya through the PERIS 2016-2020 program (SLT002/16/).

